# Circulating gut microbial metabolites and risk of coronary heart disease: a prospective, multi-stage study

**DOI:** 10.1101/2025.06.16.25329214

**Authors:** Danxia Yu, Yulu Zheng, Jae Jeong Yang, Deepak K. Gupta, David M. Herrington, Bing Yu, Ngoc Quynh H. Nguyen, Rui Pinto, Ioanna Tzoulaki, Hui Cai, Qiuyin Cai, Loren Lipworth, Xiao-Ou Shu, Wei Zheng

## Abstract

**Background:** Despite growing evidence linking gut microbiota and microbial metabolites to human cardiometabolic health, few studies have systematically examined circulating microbial metabolites with incident coronary heart disease (CHD).

**Methods:** We conducted a multi-stage metabolomics study involving five prospective cohorts. Discovery involved an untargeted plasma metabolite profiling among 896 incident cases and 896 age-/sex-/race-matched controls (∼300 pairs per race: Black, White, Asian) from the Southern Community Cohort Study (SCCS) and Shanghai Women’s Health Study and Shanghai Men’s Health Study (SWHS/SMHS). In-silico validation was conducted in the Atherosclerosis Risk in Communities Study (ARIC; N=3539; 663 cases) and Multi-Ethnic Study of Atherosclerosis (MESA; N=3860; 446 cases). Last, a quantitative assay was developed and applied to a new set of 864 cases and 864 age-/sex-/race-matched controls (∼260-340 pairs per race) from SCCS and SWHS/SMHS. Conditional logistic regression estimated odds ratios (ORs) of incident CHD per standard deviation (SD) increase in metabolite levels for discovery and quantitative stages with a nested case-control design. Cox regression was used in ARIC and MESA with a cohort design. All stages were adjusted for similar covariates.

**Results:** The discovery stage identified 73 circulating microbiota-related metabolites associated with incident CHD (FDR<0.10). Sixty-one metabolites were available for in-silico validation, with 24 showing a significant association (p<0.05) in the same direction as discovery. The targeted assay quantified eight of these 24 metabolites, with five significantly associated with incident CHD: imidazole propionate, 3-hydroxy-2-ethylpropionate, 4-hydroxyphenylacetate, trans-4-hydroxyproline, and 3-hydroxybutyrate; OR per SD ranged from 1.18 to 1.27 after adjusting for sociodemographic and lifestyle factors. The targeted assay measured eight other promising microbial metabolites, four of which were significant: trimethylamine N-oxide, phenylacetyl-L-glutamine, 4-hydroxyhippuric acid, and indolepropionate. Most associations were consistent across participant subgroups, although some (e.g., 4-hydroxyphenylacetate) were stronger among Black than White/Asian participants. Other effect modifications were found by age, obesity, and hypertension history.

**Conclusions and relevance:** We identified and validated circulating gut microbial metabolites associated with incident CHD across diverse populations. Our findings offer novel epidemiological evidence on the importance of gut microbial metabolism in CHD development and highlight specific metabolites to prioritize for mechanistic investigation, biomarker validation, and therapeutic development.

## Introduction

Coronary heart disease (CHD) remains the leading cause of death in the United States (US) and worldwide, with incidence varying by race/ethnicity, socioeconomic status, and geographic area.^1,2^ Emerging research on gut microbiota has provided insights into the etiology and prevention of CHD, with potential implications for the development of new therapeutics.^3–6^ With a genome 100 times larger than humans’, gut microbiota generates numerous small molecules (metabolites), many of which humans cannot produce. Microbial metabolites can enter host circulation and exert systemic and multifaceted effects on human health and diseases, including cardiovascular, metabolic, inflammatory, and neurological disorders.^7–10^ Prominent examples of gut microbial metabolites include short-chain fatty acids from fiber fermentation, trimethylamine N-oxide (TMAO) from phosphatidylcholine and l-carnitine, secondary bile acids from cholesterol and primary bile acids, and amino acid metabolites such as indoles from tryptophan, imidazole propionate from histidine, and phenylacetylglutamine from aromatic amino acids (AroAA).

While metabolomics has been increasingly applied to CVD research,^11–14^ evidence from human studies linking microbial metabolites to CHD or cardiovascular disease (CVD) was mainly from cross-sectional studies or clinical patient cohorts, prone to reverse causation and confounding bias. Additionally, most prior studies examined a small number of selected microbial metabolites. Comprehensive investigations of microbial metabolites in population-based, prospective studies with rigorous validation of findings are crucial. Further, few prospective studies examined microbial metabolites in populations with high CVD burden,^15,16^ such as Black Americans and low-income individuals. For gut microbiota-related research, data from populations varying in sociodemographic, geographic, and diet/lifestyle backgrounds hold significant value. Studies have shown distinct metabolomic profiles among individuals with different diets/lifestyles, with many distinguishing metabolites related to gut microbiota.^17–19^ Whether microbial metabolites contribute to CHD risk across diverse populations warrants investigation.

Hence, we conducted a multi-stage metabolomic study involving demographically and geographically diverse participants, including 1) an untargeted, semi-quantitative assay for discovery among 896 incident CHD cases and 896 age-/sex-/race-matched controls (∼300 pairs per race: Black, White, and Asian/Chinese) from the Southern Community Cohort Study (SCCS) and Shanghai Women’s Health Study and Shanghai Men’s Health Study (SWHS/SMHS); 2) an in-silico validation in the Atherosclerosis Risk in Communities Study (ARIC) and Multi-Ethnic Study of Atherosclerosis (MESA); 3) a targeted, quantitative assay to measure metabolite concentrations, verify their associations with CHD, and evaluate potential microbiota-host interactions in a new set of 864 incident CHD cases and 864 matched controls (∼260-340 pairs per race) from SCCS and SWHS/SMHS. This study aimed to enhance the understanding of the role of gut microbial metabolites in CHD development and inform potential novel biomarkers or prevention strategies.

## Methods

### Study population and design

Detailed descriptions of the study designs and protocols for SCCS, SWHS, SMHS, ARIC, and MESA were published elsewhere.^20–25^ Briefly, all are population-based prospective cohorts that recruited participants, conducted surveys, collected biospecimens, including peripheral blood, and followed participants for disease outcomes, including incident CHD and CHD death. SCCS enrolled ∼85,000 participants (40–79 years) from 12 southern US states in 2002–2009, mostly Black/African American adults with low household incomes.^20^ SWHS enrolled ∼75,000 women (40–70 years), and SMHS enrolled ∼61,000 men (40–74 years) from Shanghai, China, in 1996-2000 and 2002-2006, respectively.^21,22^ ARIC enrolled ∼16,000 participants (45–65 years) from four US centers in 1987–1989, with ∼27% being Black/African American.^23,24^ MESA enrolled 6,814 participants (45–84 years) from six US centers in 2000–2002, with ∼38% being White, 28% Black/African American, 22% Hispanic/Latino, and 12% Chinese.^25^ See **Supplementary Methods** for detailed information. All cohorts received IRB approval from the participating institutions. The Vanderbilt University Medical Center IRB approved the present study.

For discovery and targeted validation, nested case-control studies were conducted in SCCS and SWHS/SMHS. Participant inclusion criteria were 1) no history of CHD, stroke, heart failure, cancer, or end-stage renal disease at baseline; 2) available plasma samples and data on fasting time, and in SCCS, the time between sample collection and lab processing; 3) no use of antibiotics nor cold/flu in the last seven days before blood collection to minimize the potential influence of acute disease and medications on gut microbiota; 4) in SCCS, participants were covered by the Centers for Medicare & Medicaid Services (CMS) and had ≥2 claims after cohort enrollment through December 2016; in SWHS/SMHS, participants’ medical records were accessible for this study through December 2016. In SCCS, nonfatal CHD cases were identified through CMS data, including acute myocardial infarction (MI) and coronary revascularization, and CHD deaths were identified through the National Death Index. In SWHS/SMHS, CHD cases were initially identified by self-reported physician diagnoses during follow-up visits and confirmed by medical records. CHD cases were 1:1 matched with controls without a history of CVD or cancer at the time of case diagnosis using incidence density sampling by race, sex, enrollment age (±2 years), fasting time (±2 hours), and time between blood collection and lab processing (±4 hours for SCCS; all SWHS/SMHS samples were processed within 6 hours after collection). After applying inclusion and case-control matching criteria, 150 case-control pairs in each race (Black, White, Asian) and sex (male, female) were randomly selected for discovery, with the reminder included for targeted validation: Black women (167 pairs), Black men (170 pairs), White women (149 pairs), White men (109 pairs), Chinese women (120 pairs), and Chinese men (148 pairs).

The in-silico validation was conducted in ARIC and MESA using a cohort design. In ARIC, 3,539 participants with plasma metabolite data at visit 1 were included after excluding those with a history of CHD, heart failure, stroke, or cancer. Incident CHD was defined as definite or probable MI, fatal CHD, and if participants had undergone any cardiac procedures or ECG MI before December 31, 2018. In MESA, 3,860 participants with baseline plasma metabolite data were included after excluding participants with a history of heart disease or stroke or not fasting. Incident CHD was defined as MI, resuscitated cardiac arrest, and CHD death occurring on or before and adjudicated through December 31, 2018.

### Metabolites profiling

For discovery and targeted validation, baseline plasma samples of the case-control pairs were retrieved and placed adjacent in the same batch at random. Laboratory personnel were blinded to the case-control status. For discovery, untargeted metabolite profiling was performed using ultra-high-performance liquid chromatography with tandem mass spectrometry (UHPLC-MS/MS) in positive and negative ion modes using a combination of reverse phase and HILIC chromatography methods by Metabolon Inc. The general assay protocol has been published.^26^ A brief description is provided in **Supplementary Methods**. Overall, plasma samples were extracted with methanol and split into four aliquots for UHPLC-MS/MS assays in both positive and negative ion modes using a combination of reverse phase and HILIC chromatography methods. Metabolites were identified by comparing mass spectral features to a reference library of >5,000 authenticated standard compounds. A total of 1503 metabolites were detected in our discovery stage samples. For the present study, we focused on 226 gut microbiota-related metabolites identified by two methods: 1) linkage to a database of 460 confirmed gut microbial metabolites via Exposome-Explorer;^27^ 2) regression analysis of metabolite levels with oral antibiotic use seven days before blood collection to identify significantly reduced metabolites using previous data from SWHS/SMHS (1,841 participants; 1,498 metabolites measured by the same Metabolon panel). These two methods identified 165 and 61 microbiota-related metabolites, respectively. The median coefficient of variation (CV) for those 226 metabolites among our QC samples was 9.7% (25^th^-75^th^ percentile: 5.6%-16.3%).

In ARIC, baseline fasting serum samples were analyzed by the same Metabolon panel using UHPLC-MS/MS, which detected 787 metabolites.^26^ In MESA, baseline samples were analyzed using nuclear magnetic resonance (NMR) and LC-MS, with detailed methods previously published.^28,29,30^ A brief description is provided in **Supplementary Methods.**

For targeted validation, quantitative measurements of metabolites were performed using LC-MS/MS. Specifically, plasma samples were spiked with isotopically labeled internal standards and subjected to protein precipitation with an acidified organic solution. An aliquot was injected onto an Agilent 1290/Sciex Triple Quad 6500+ LC-MS/MS system equipped with either an Agilent Zorbax SB-C18 RRHD for the reverse phase method or a Waters Acquity Premier BEH Amide column for the HILIC method. The mass spectrometer was operated in both positive and negative modes using heated electrospray ionization. The peak area of the individual analyte product ions was measured against the peak area of the product ions of the corresponding internal standards. Quantitation was performed using a weighted linear least squares regression analysis generated from fortified calibration standards prepared concurrently with study samples. Sample analysis was conducted in a 96-well plate format containing two calibration curves and six QC samples (three levels with two at each). Accuracy was evaluated using the QC replicates in the sample runs. The average accuracy for QC at all levels for all 16 analytes was >90% except for 1-methyl-4-imidazoleacetate (88%). The median CV for the 16 metabolites among our study QC samples was 3.6% (25^th^-75^th^ percentile: 1.6%-6.4%). Detailed information on reference internal standards, calibration ranges, and QC performance can be found in **Supplementary Methods**.

Within each study and each batch, metabolite levels below the detection limit were imputed using the minimum detection value, then log-transformed and z-scored prior to statistical analyses.

### Statistical analysis

In discovery and targeted validation stages with a matched case-control design, conditional logistic regression was applied to compute odds ratio (OR) and 95% confidence interval (95% CI) for a standard deviation (SD) increase in log-transformed metabolites among all participants and by race. Model 1 adjusted for age (continuous); Model 2 additionally adjusted for educational attainment (less than high school, high school graduation, vocational school, college or more), income (cohort-specific low, middle, and high), smoking status (never, former, current with <20 cigarettes/day, current with ≥20 cigarettes/day), alcohol consumption (none, moderate [≤2 drinks/day for men, ≤1 drink/day for women], heavy), physical activity (cohort-specific tertiles), diet quality score (cohort-specific tertiles), and body mass index (BMI) (continuous). We also conducted stratified analyses by baseline status of diabetes, hypertension, and dyslipidemia using logistic regression with additional adjustment for matching variables (sex, race, fasting status). Metabolites showing Benjamini–Hochberg false discovery rate (FDR) adjusted p-values<0.1 in either model among all participants or in any subgroups by race/disease status entered in-silico validation. The in-silico validation was conducted in ARIC and MESA using Cox regression with the same covariates described above, plus center and batch. The analyses were performed within each cohort among all eligible participants and by race and baseline diabetes, hypertension, and dyslipidemia. Metabolites were considered validated if nominal p<0.05 in any cohort with the same direction of association. SAS Enterprise (SAS Institute, Cary, North Carolina, US) and R (version 4.3.3) were used for analyses.

## Results

### Baseline characteristics of study participants

Mean age at baseline was ∼57 years (SD: 9) for included SCCS and SWHS/SMHS participants (**Table 1**). Compared to age/sex/race-matched controls, individuals who developed incident CHD over a mean follow-up of ∼6 years (SD: 4) had lower levels of education and household income and were more likely to be current smokers, non-drinkers, and have low leisure-time physical activity. In addition, CHD cases showed higher baseline BMI and greater prevalence of diabetes, hypertension, dyslipidemia, and family history of CHD. Characteristics of participants included in in-silico validation are shown in **Supplementary Table 1**. Briefly, ARIC participants had a mean age of ∼54 years at visit 1, with 60.3% being women, 38.5% White, and 61.5% Black/African American. MESA participants had a mean age of ∼63 years at baseline, with 50.7% being women, 39.1% non-Hispanic White, 23.6% Black/African American, 13.7% Chinese American, and 23.7% Hispanic.

**Table 1.**
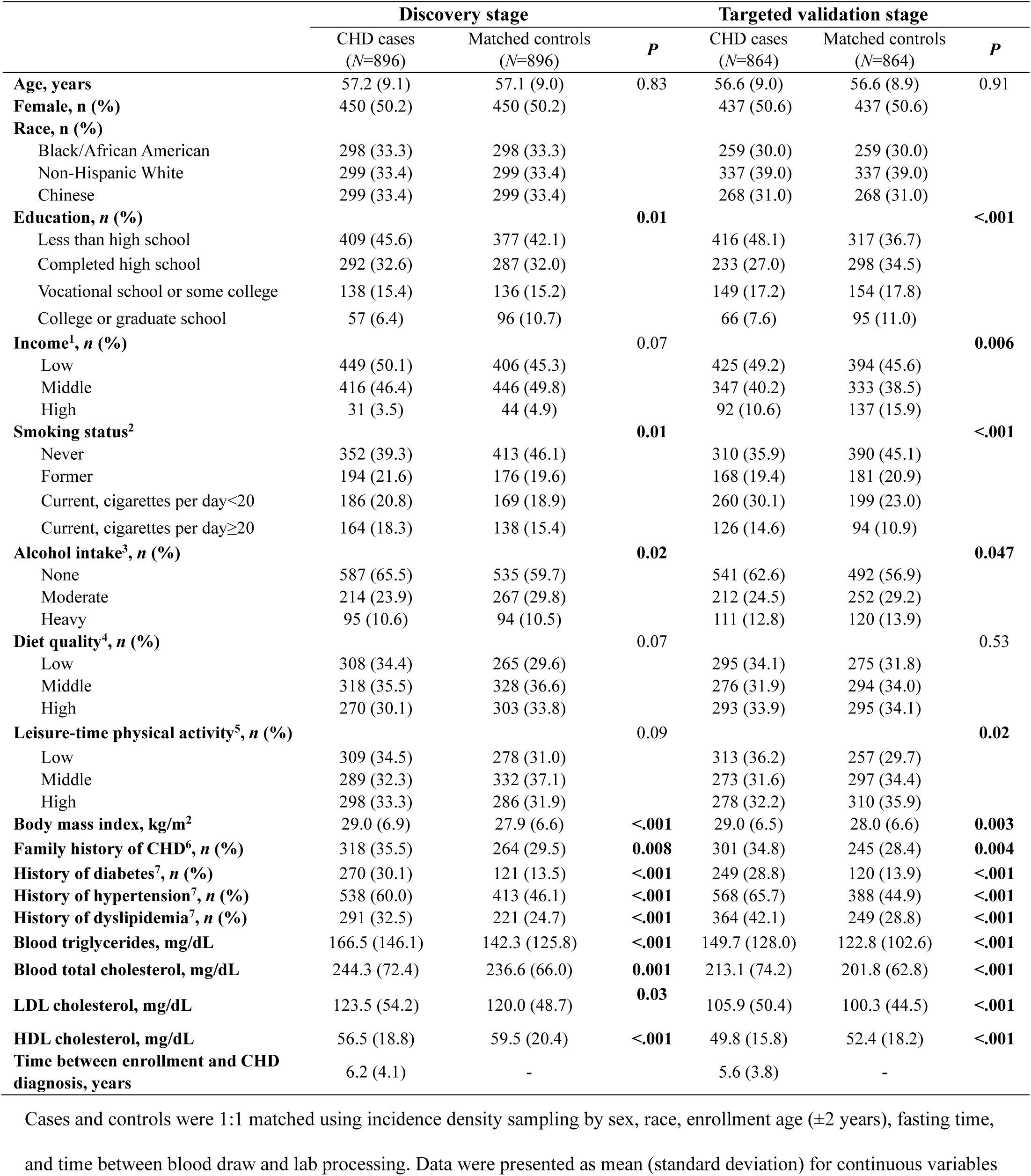
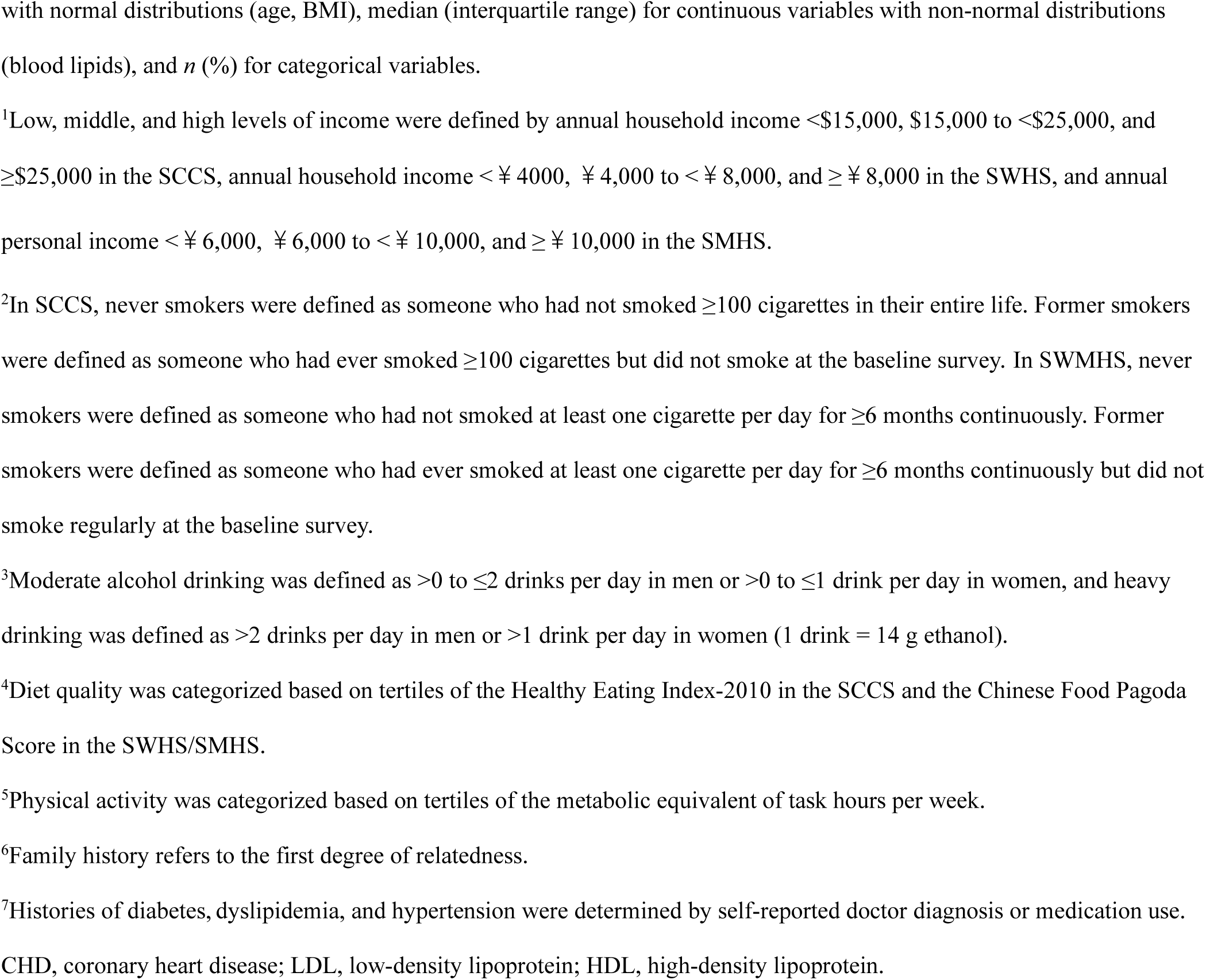
Baseline characteristics of participants in the discovery and targeted validation stages.

|  | Discovery stage |  |  | Targeted validation stage |  |  |
| --- | --- | --- | --- | --- | --- | --- |
|  | CHD cases<br>(N=896) | Matched controls<br>(N=896) | P | CHD cases<br>(N=864) | Matched controls<br>(N=864) | P |
| **Age, years** | 57.2 (9.1) | 57.1 (9.0) | 0.83 | 56.6 (9.0) | 56.6 (8.9) | 0.91 |
| **Female, n (%)** | 450 (50.2) | 450 (50.2) |  | 437 (50.6) | 437 (50.6) |  |
| **Race, n (%)** |  |  |  |  |  |  |
| Black/African American | 298 (33.3) | 298 (33.3) |  | 259 (30.0) | 259 (30.0) |  |
| Non-Hispanic White | 299 (33.4) | 299 (33.4) |  | 337 (39.0) | 337 (39.0) |  |
| Chinese | 299 (33.4) | 299 (33.4) |  | 268 (31.0) | 268 (31.0) |  |
| **Education, n (%)** |  |  | **0.01** |  |  | **<.001** |
| Less than high school | 409 (45.6) | 377 (42.1) |  | 416 (48.1) | 317 (36.7) |  |
| Completed high school | 292 (32.6) | 287 (32.0) |  | 233 (27.0) | 298 (34.5) |  |
| Vocational school or some college | 138 (15.4) | 136 (15.2) |  | 149 (17.2) | 154 (17.8) |  |
| College or graduate school | 57 (6.4) | 96 (10.7) |  | 66 (7.6) | 95 (11.0) |  |
| **Income<sup>1</sup>, n (%)** |  |  | 0.07 |  |  | **0.006** |
| Low | 449 (50.1) | 406 (45.3) |  | 425 (49.2) | 394 (45.6) |  |
| Middle | 416 (46.4) | 446 (49.8) |  | 347 (40.2) | 333 (38.5) |  |
| High | 31 (3.5) | 44 (4.9) |  | 92 (10.6) | 137 (15.9) |  |
| **Smoking status<sup>2</sup>** |  |  | **0.01** |  |  | **<.001** |
| Never | 352 (39.3) | 413 (46.1) |  | 310 (35.9) | 390 (45.1) |  |
| Former | 194 (21.6) | 176 (19.6) |  | 168 (19.4) | 181 (20.9) |  |
| Current, cigarettes per day<20 | 186 (20.8) | 169 (18.9) |  | 260 (30.1) | 199 (23.0) |  |
| Current, cigarettes per day≥20 | 164 (18.3) | 138 (15.4) |  | 126 (14.6) | 94 (10.9) |  |
| **Alcohol intake<sup>3</sup>, n (%)** |  |  | **0.02** |  |  | **0.047** |
| None | 587 (65.5) | 535 (59.7) |  | 541 (62.6) | 492 (56.9) |  |
| Moderate | 214 (23.9) | 267 (29.8) |  | 212 (24.5) | 252 (29.2) |  |
| Heavy | 95 (10.6) | 94 (10.5) |  | 111 (12.8) | 120 (13.9) |  |
| **Diet quality<sup>4</sup>, n (%)** |  |  | 0.07 |  |  | 0.53 |
| Low | 308 (34.4) | 265 (29.6) |  | 295 (34.1) | 275 (31.8) |  |
| Middle | 318 (35.5) | 328 (36.6) |  | 276 (31.9) | 294 (34.0) |  |
| High | 270 (30.1) | 303 (33.8) |  | 293 (33.9) | 295 (34.1) |  |
| **Leisure-time physical activity<sup>5</sup>, n (%)** |  |  | 0.09 |  |  | **0.02** |
| Low | 309 (34.5) | 278 (31.0) |  | 313 (36.2) | 257 (29.7) |  |
| Middle | 289 (32.3) | 332 (37.1) |  | 273 (31.6) | 297 (34.4) |  |
| High | 298 (33.3) | 286 (31.9) |  | 278 (32.2) | 310 (35.9) |  |
| **Body mass index, kg/m<sup>2</sup>** | 29.0 (6.9) | 27.9 (6.6) | **<.001** | 29.0 (6.5) | 28.0 (6.6) | **0.003** |
| **Family history of CHD<sup>6</sup>, n (%)** | 318 (35.5) | 264 (29.5) | **0.008** | 301 (34.8) | 245 (28.4) | **0.004** |
| **History of diabetes<sup>7</sup>, n (%)** | 270 (30.1) | 121 (13.5) | **<.001** | 249 (28.8) | 120 (13.9) | **<.001** |
| **History of hypertension<sup>7</sup>, n (%)** | 538 (60.0) | 413 (46.1) | **<.001** | 568 (65.7) | 388 (44.9) | **<.001** |
| **History of dyslipidemia<sup>7</sup>, n (%)** | 291 (32.5) | 221 (24.7) | **<.001** | 364 (42.1) | 249 (28.8) | **<.001** |
| **Blood triglycerides, mg/dL** | 166.5 (146.1) | 142.3 (125.8) | **<.001** | 149.7 (128.0) | 122.8 (102.6) | **<.001** |
| **Blood total cholesterol, mg/dL** | 244.3 (72.4) | 236.6 (66.0) | **0.001** | 213.1 (74.2) | 201.8 (62.8) | **<.001** |
| **LDL cholesterol, mg/dL** | 123.5 (54.2) | 120.0 (48.7) | **0.03** | 105.9 (50.4) | 100.3 (44.5) | **<.001** |
| **HDL cholesterol, mg/dL** | 56.5 (18.8) | 59.5 (20.4) | **<.001** | 49.8 (15.8) | 52.4 (18.2) | **<.001** |
| **Time between enrollment and CHD diagnosis, years** | 6.2 (4.1) | - |  | 5.6 (3.8) | - |  |

Cases and controls were 1:1 matched using incidence density sampling by sex, race, enrollment age ( $\pm 2$ years), fasting time, and time between blood draw and lab processing. Data were presented as mean (standard deviation) for continuous variables
with normal distributions (age, BMI), median (interquartile range) for continuous variables with non-normal distributions (blood lipids), and *n* (%) for categorical variables.
<sup>1</sup>Low, middle, and high levels of income were defined by annual household income <\$15,000, \$15,000 to <\$25,000, and ≥\$25,000 in the SCCS, annual household income <¥ 4000, ¥ 4,000 to <¥ 8,000, and ≥ ¥ 8,000 in the SWHS, and annual personal income < ¥ 6,000, ¥ 6,000 to < ¥ 10,000, and ≥ ¥ 10,000 in the SMHS.
<sup>2</sup>In SCCS, never smokers were defined as someone who had not smoked ≥100 cigarettes in their entire life. Former smokers were defined as someone who had ever smoked ≥100 cigarettes but did not smoke at the baseline survey. In SWMHS, never smokers were defined as someone who had not smoked at least one cigarette per day for ≥6 months continuously. Former smokers were defined as someone who had ever smoked at least one cigarette per day for ≥6 months continuously but did not smoke regularly at the baseline survey.
<sup>3</sup>Moderate alcohol drinking was defined as >0 to ≤2 drinks per day in men or >0 to ≤1 drink per day in women, and heavy drinking was defined as >2 drinks per day in men or >1 drink per day in women (1 drink = 14 g ethanol).
<sup>4</sup>Diet quality was categorized based on tertiles of the Healthy Eating Index-2010 in the SCCS and the Chinese Food Pagoda Score in the SWHS/SMHS.
<sup>5</sup>Physical activity was categorized based on tertiles of the metabolic equivalent of task hours per week.
<sup>6</sup>Family history refers to the first degree of relatedness.
<sup>7</sup>Histories of diabetes, dyslipidemia, and hypertension were determined by self-reported doctor diagnosis or medication use. CHD, coronary heart disease; LDL, low-density lipoprotein; HDL, high-density lipoprotein.

### Discovery of circulating microbiota-related metabolites associated with incident CHD

In the discovery stage, 48 out of 226 microbiota-related metabolites showed a significant association with incident CHD (FDR<0.1) in Model 1. All but five metabolites remained significant after further adjustments for sociodemographics, lifestyles, BMI, and prevalent metabolic diseases (FDR<0.1 in Model 2); meanwhile, two metabolites became significant after additional adjustments. Furthermore, 23 metabolites were significantly associated with incident CHD in participant subgroups, including 19 in race-specific analyses and 4 in stratified analyses by metabolic disease status. A total of 73 metabolites were significant in either model among all participants or subgroups by race/disease status. They were from super pathways of amino acids (n=28), lipids (n=23), nucleotides (n=5), carbohydrates (n=4), energy (n=4), xenobiotics (n=4), cofactors/vitamins (n=3), and unknowns (n=2), with ORs per SD ranging from 0.61 to 0.88 for inverse associations and 1.13 to 1.71 for positive associations (**Fig. 1**). Detailed information (e.g., sub-pathway, HMDB ID, PubChem ID) and ORs (95% CI) for those metabolites in total participants and subgroups are in **Supplementary Table 2**.

**Fig. 1.**
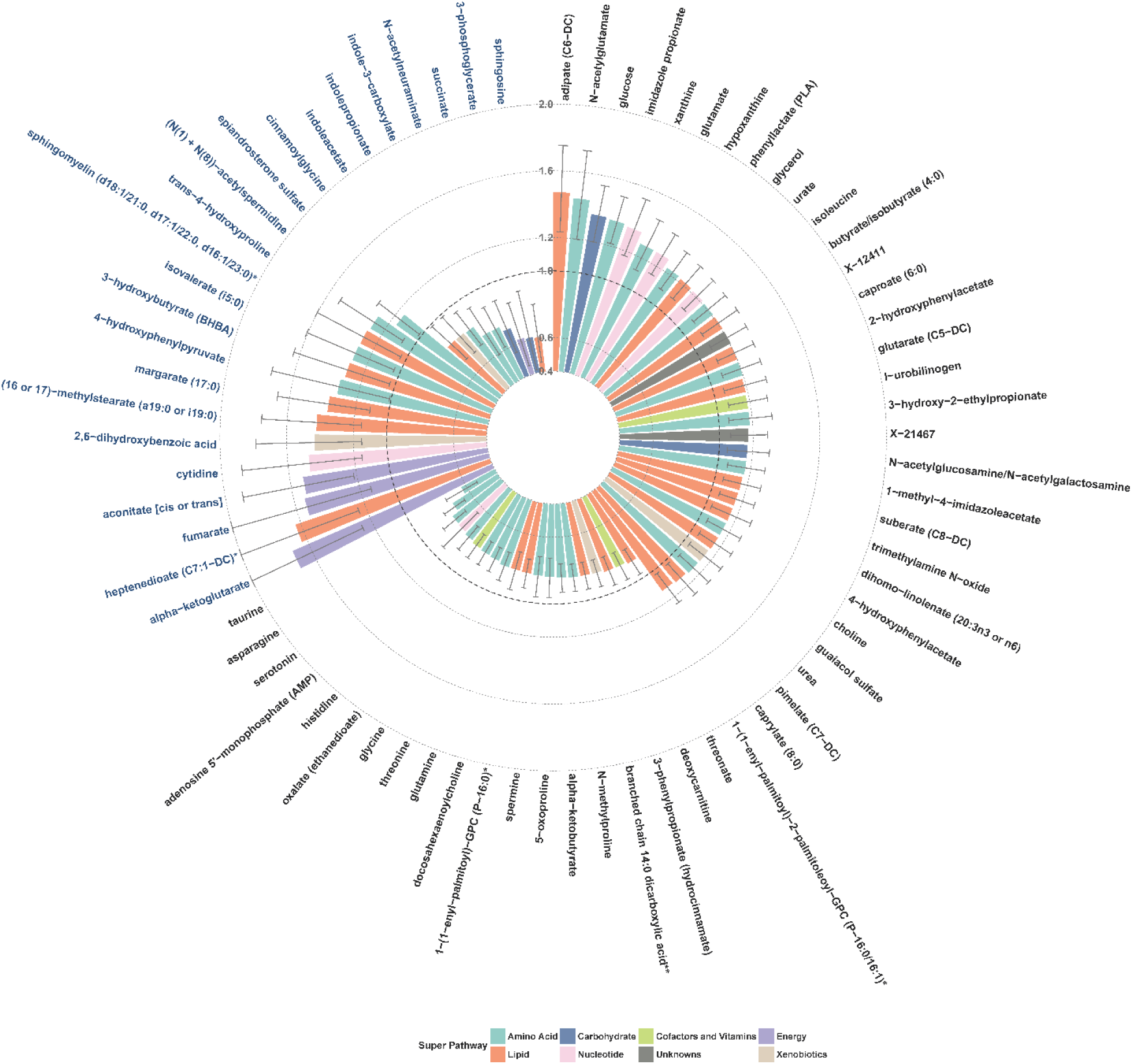
Discovery of circulating gut microbiota-related metabolites associated with incident coronary heart disease. The discovery stage applied untargeted, semi-quantitative metabolite profiling among 896 incident cases and 896 matched controls from SCCS and SWHS/SMHS. ORs and 95% CIs for per SD increase in log-transformed metabolite levels were obtained using conditional logistic regression among all participants and by race or logistic regression by metabolic disease status with adjustment for matching variables (age, sex, race, and fasting status). Metabolites showing FDR-p<0.1 among total participants (black font) or in participant subgroups (blue font) are presented.

### In-silico validation of circulating microbiota-related metabolites associated with incident CHD

Among the 73 metabolites, 61 were available in ARIC or MESA, and 24 showed a significant association (p<0.05 in either cohort) in the same direction as discovery (**Fig. 2**). Most validated metabolites were from super pathways of amino acids (n=17), including metabolites of histidine, glutamate, phenylalanine, tyrosine, isoleucine, arginine and proline, glycine, and tryptophan. Others included metabolites of lipids and xenobiotics.

**Fig. 2.**
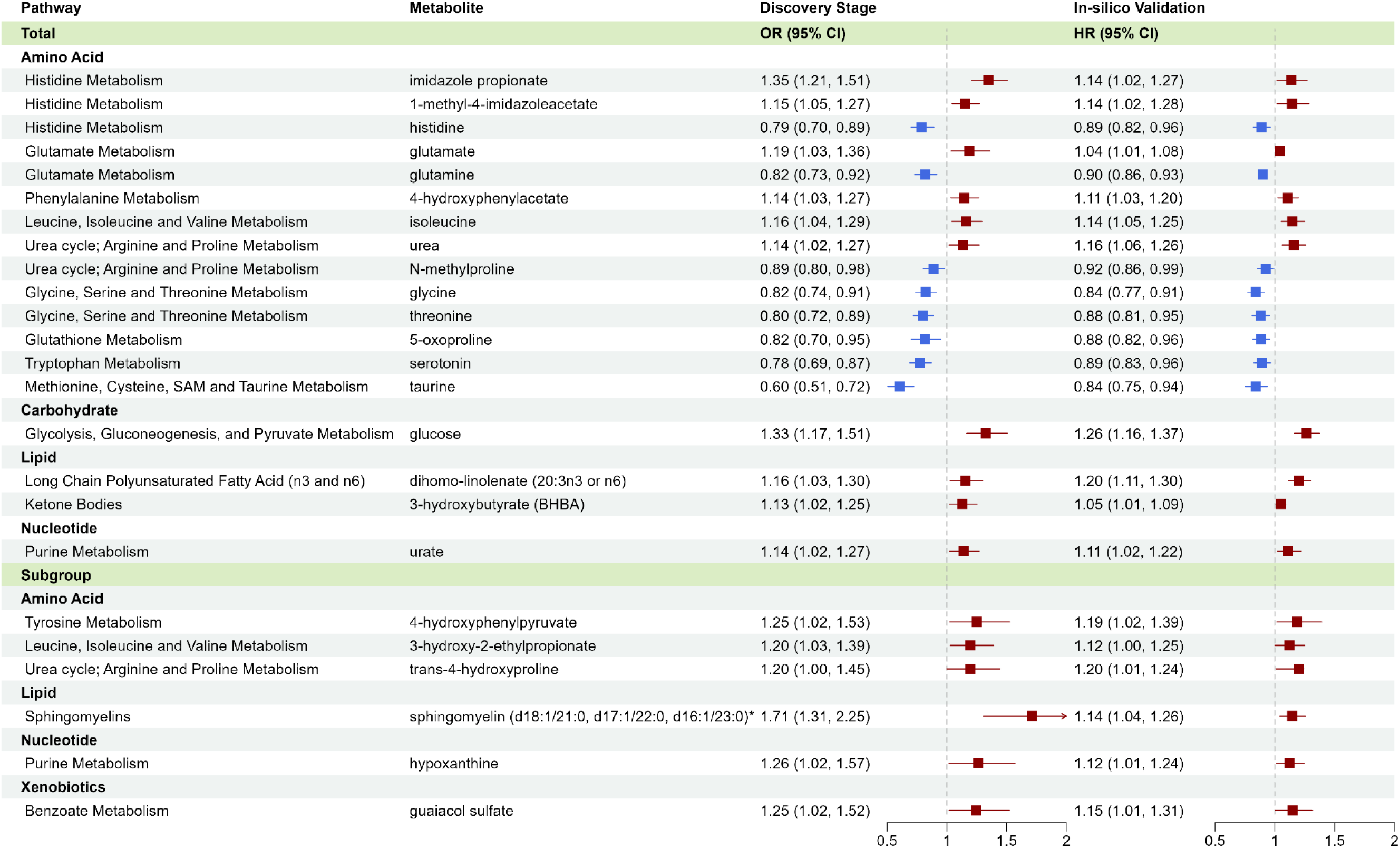
In-silico validation of circulating gut microbiota-related metabolites associated with incident coronary heart disease. The in-silico validation stage was conducted using existing untargeted, semi-quantitative metabolite data among 3,539 participants in ARIC (663 cases) and 3,860 participants in MESA (446 cases). HRs and 95% CIs for per SD increase in log-transformed metabolite levels were obtained using Cox regression among all participants and by race or metabolic disease status with adjustments for sociodemographics, lifestyles, and BMI. Metabolites showing p<0.05 in the same direction as the discovery results among total participants or participant subgroups are presented.

### Targeted validation of circulating microbiota-related metabolites associated with incident CHD

A quantitative assay was designed to measure concentrations of the most promising metabolites, which captured eight of 24 metabolites that passed the in-silico validation, including seven amino acid metabolites (imidazole-propionate, 1-methyl-4-imidazoleacetate, 3-hydroxy-2-ethylpropionate, 4-hydroxyphenylacetate, trans-4-hydroxyproline, 4-hydroxyphenylpyruvate, taurine) and ketone body 3-hydroxybutyrate. Due to different LC-MS requirements, the remaining 16 metabolites could not be quantified simultaneously. However, the assay was able to measure eight other metabolites that were significant in the discovery and reported in recent population studies linking them to CHD/CVD but unavailable or insignificant in in-silico validation, including metabolites of amino acids (phenylacetyl-L-glutamine, alpha-ketobutyrate, indolepropionate), lipids (TMAO), and xenobiotics (3-phenylpropionate, 4-hydroxyhippuric acid, 2,6-dihydroxybenzoic acid, p-cresol-sulfate). Plasma concentrations of the 16 metabolites among cases and controls and distributions by race are shown in **Supplementary Table 3 and Supplementary Figure**. After adjusting for sociodemographics and lifestyles, nine metabolites were significant (**Figure 3**; p<0.05 in Model 2), including 3-hydroxybutyrate (OR [95% CI] per SD increase = 1.27 [1.14-1.42]), imidazole-propionate (1.26 [1.11-1.42]), 3-hydroxy-2-ethylpropionate (1.24 [1.09-1.41]), TMAO (1.22 [1.09-1.36]), 4-hydroxyphenylacetate (1.19 [1.06-1.33]), phenylacetyl-L-glutamine (1.14 [1.02-1.27]), trans-4-hydroxyproline (1.18 [1.04-1.35]), 4-hydroxyhippuric acid (1.18 [1.05-1.32]), and indolepropionate (0.89 [0.80-0.99]). Most associations remained significant after further adjustment for baseline diabetes, hypertension, and dyslipidemia (Model 3), except for 3-hydroxy-2-ethylpropionate, phenylacetyl-L-glutamine, and indolepropionate, suggesting they may affect incident CHD through those conditions.

**Fig. 3.**
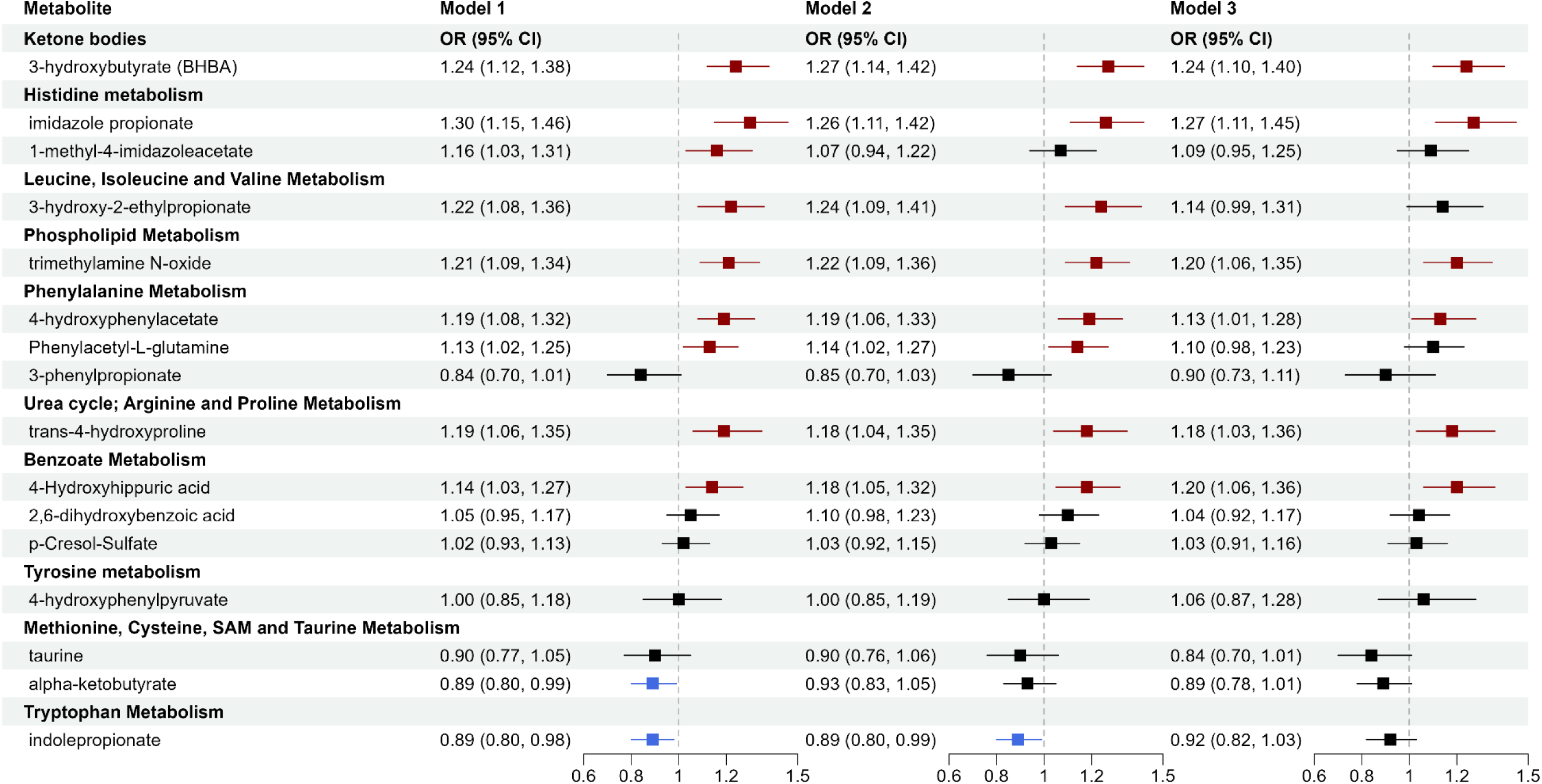
Targeted validation of circulating gut microbiota-related metabolites associated with incident coronary heart disease. The targeted validation stage applied quantitative profiling and measured 16 metabolites among 864 incident cases and 864 matched controls from SCCS and SWHS/SMHS. ORs and 95% CIs for per SD increase in log-transformed metabolite levels were obtained using conditional logistic regression among all participants and by race or logistic regression by metabolic disease status with adjustment for matching variables (age, sex, race, and fasting status). Model 1 adjusted for age (continuous). Model 2 additionally adjusted for education, income, tobacco smoking, alcohol consumption, physical activity, diet quality score, and BMI (continuous). Model 3 additionally adjusted for history of diabetes, hypertension, and dyslipidemia.

We further evaluated whether those metabolite-CHD associations differ by participant demographics, lifestyles, metabolic disease history, and follow-up time through stratified analyses and interaction testing. Several metabolites showed stronger associations among Black than White or Asian participants (**Fig. 4**), including 4-hydroxyphenylacetate, phenylacetyl-L-glutamine, 3-hydroxy-2-ethylpropionate, and trans-4-hydroxyproline (ORs per SD =1.37, 1.32, 1.29, and 1.26 among Black participants, respectively, with p-interaction<0.05 for the first two). Meanwhile, 1-methyl-4-imidazoleacetate was significant only among White participants (OR=1.73; p-interaction<0.05). TMAO was significant only among Americans (OR=1.39 among Black and 1.45 among White participants). Other potential effect modifications include 4-hydroxyhippuric acid by age (stronger in the older group) and 3-hydroxy-2-ethylpropionate by obesity and hypertension status (stronger among those without obesity or hypertension). P-cresol-sulfate and 3-phenylpropionate were only significant for CHD diagnosed within two years after baseline. No significant interactions were found by status of diabetes or dyslipidemia, physical activity, or diet quality. See detailed results in **Supplementary Table 4**.

**Fig. 4.**
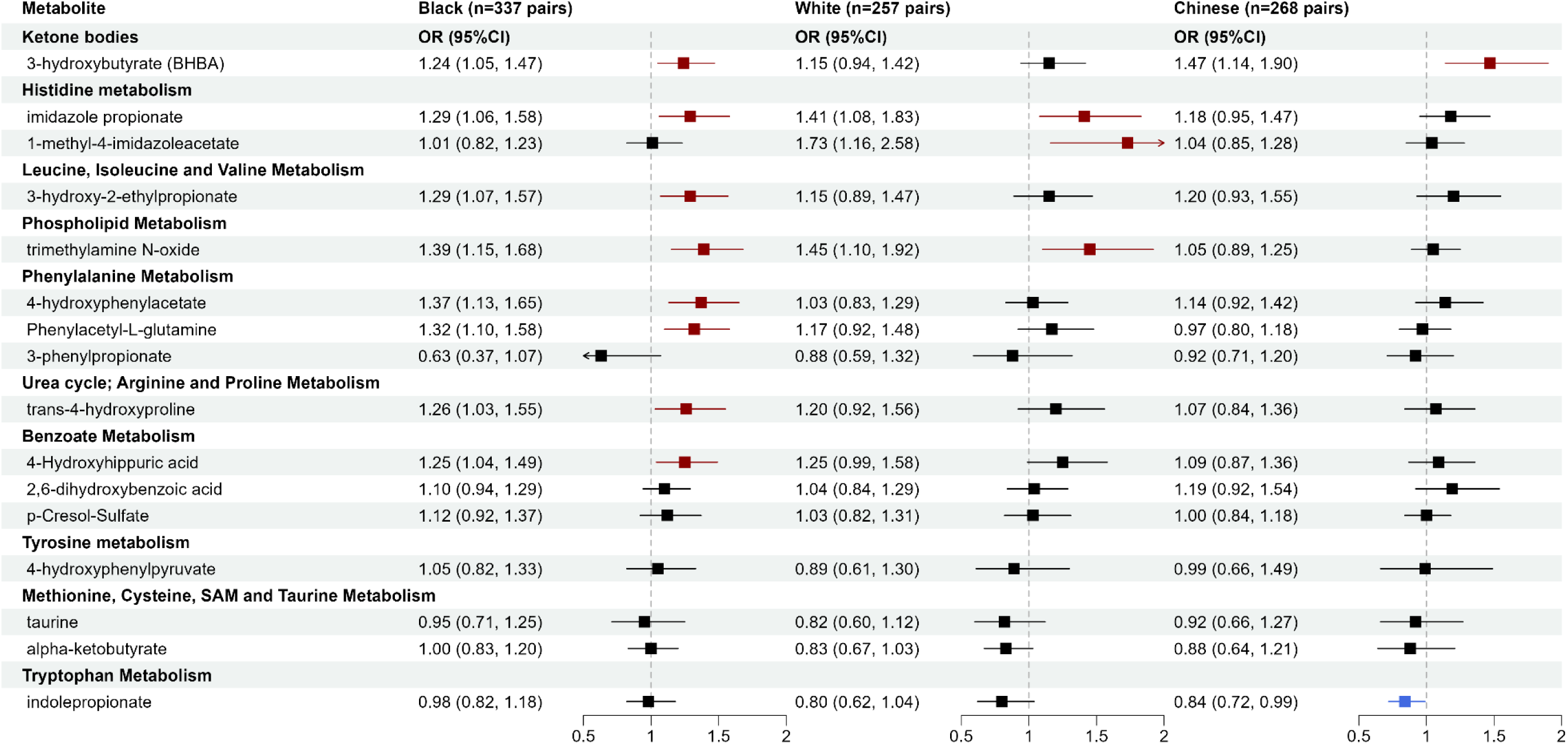
Circulating gut microbiota-related metabolites associated with incident coronary heart disease by race. ORs and 95% CIs for per SD increase in log-transformed metabolite levels were obtained using conditional logistic regression in each racial group, adjusting for age, education, income, tobacco smoking, alcohol consumption, physical activity, diet quality score, and BMI (Model 2).

## Discussion

In this multi-stage metabolomic study involving five prospective cohorts, we identified and validated circulating gut microbiota-related metabolites linked to incident CHD. Major strengths include demographically and geographically diverse participants and a rigorous design with discovery, in-silico validation, and quantitative validation. Most of the significant findings were consistent across racial groups and participant metabolic health statuses, suggesting broad generalizability of those metabolite-CHD associations. This study presents new epidemiological evidence supporting the role of gut microbiota in CHD etiology and highlights microbial metabolites and pathways as potential novel biomarkers or therapeutic targets for future mechanistic and interventional studies.

### Microbial metabolites of amino acids with incident CHD

Our results, particularly from quantitative validation, confirmed the role of microbial amino acid metabolites in cardiometabolic diseases. Notably, microbial metabolites of AroAA (phenylalanine, tyrosine, tryptophan) were linked to CVD risk in recent large studies, including the MetaCardis consortium of 1241 European adults,^14^ Cleveland Clinic GeneBank and LipidCardio study of 4000 US and 833 European adults undergoing elective coronary angiography,^31^ and 7897 US and European adults from 6 cohorts, including ARIC, in the COnsortium of METabolomics Studies (COMETS).^13^ Specifically, phenylalanine metabolites hydroxyphenylacetate and phenylacetyl-L-glutamine were associated with increased risk of CHD or major adverse cardiovascular events (MACE) in all those studies. Moreover, effect sizes were similar: HR for 4-hydroxyphenylacetate was 1.24 in COMETS (OR=1.19 in ours); HR for phenylacetyl-L-glutamine was 1.12 in COMETS and 1.17 among 4241 Swedish adults from the Malmö cohorts^32^ (OR=1.14 in ours). Among tyrosine metabolites, 4-hydroxyphenylpyruvate was significant among participants with dyslipidemia in discovery and in-silico validation but failed quantitative validation. However, 4-hydroxyhippuric acid, a tyrosine pathway and benzoate metabolite, was significant in quantitative stage (OR=1.20 even after adjustment for metabolic diseases), but it was not measured in discovery stage. We added 4-hydroxyhippuric acid to the quantitative assay because of its association with incident MI/MACE in recent studies^31^ and confirmed this. Conversely, tryptophan metabolite indolepropionate was associated with lower CHD risk in discovery and quantitative stages, although it was insignificant in ARIC and unavailable in MESA for in-silico validation. Its association attenuated after adjusting for prevalent diabetes (OR=0.89 in Model 2 to 0.92 in Model 3), consistent with evidence on its major role in glucose metabolism and diabetes prevention.^33–35^

Besides AroAA metabolites, our study elucidated other amino acid metabolites, including imidazole-propionate (from histidine) and 3-hydroxy-2-ethylpropionate (from isoleucine). The EPIC-Norfolk cohort first reported circulating imidazole-propionate linked to incident CHD and other diseases (heart failure, renal disease), adjusted for age and sex.^12^ Our study confirmed and extended its association with incident CHD in a diverse population, which remained significant after adjusting for sociodemographics, lifestyle, and metabolic diseases (p<0.05 at all stages with ORs=1.35, 1.11, and 1.26). 3-hydroxy-2-ethylpropionate (also named 2-ethylhydracrylic acid) indicates isoleucine catabolism defects.^36^ Despite substantial evidence linking circulating branched-chain amino acids (BCAA: valine, leucine, isoleucine) with cardiometabolic diseases,^37–39^ to our knowledge, no studies have reported elevated circulating 3-hydroxy-2-ethylpropionate with incident CHD. Its precursor, isoleucine, was also elevated in baseline plasma of incident CHD cases (Fig. 2). Together, those findings support that impaired BCAA catabolism contributes to CHD development.

### Microbiota-related metabolites of lipids with incident CHD

Our study also linked microbial metabolites from the lipid pathway to incident CHD, including TMAO (from choline, phosphatidylcholine, and l-carnitine), 3-hydroxybutyrate (from fatty acids and ketogenic amino acids), and sphingomyelin among participants with dyslipidemia. TMAO exemplifies how gut microbial metabolism of dietary intakes (e.g., egg and red meat) affects CVD risk and prognosis.^40,41^ Our previous cross-sectional analysis of >32,000 adults from the US, Europe, and Asia reported that elevated circulating TMAO correlated with high animal food intakes and CVD risk factors, including impaired renal function and glycemic control.^42^ Our current prospective analysis confirmed its link with incident CHD (Fig. 3, OR∼1.20), particularly among Americans (Fig. 4, OR∼1.40). On the other hand, the role of ketone bodies in cardiometabolic health is increasingly discussed but remains controversial. We observed a positive association between circulating 3-hydroxybutyrate, the primary ketone body, and incident CHD at all stages (OR=1.24 in model 3 of quantitative stage), particularly among individuals without obesity (OR=1.36). This finding seemed contradictory to emerging research on ketogenic diets or supplements^43^ to improve cardiometabolic health. Among free-living individuals without intervention, elevated circulating ketones are more likely due to decreased ketone oxidation rather than increased ketogenesis. Nevertheless, given our significant positive association and inconsistent findings from clinical trials on ketogenic diets/supplements, further research is warranted to clarify the role of ketone bodies/signaling in CVD prevention.

### Strengths and limitations

The major strengths of our study include a diverse population, multi-stage design, and quantitative validation. Analyses by race and metabolic diseases indicated that most metabolite-CHD associations were consistent across participant groups. Quantitative assay revealed that concentrations of many metabolites were comparable between Black and White Americans but differed between US and Asian adults. We presented detailed results, including metabolite IDs, concentrations, and subgroup associations, in the Supplementary files to inform future research. Meanwhile, this is the first study to identify and validate circulating microbiota-related metabolites with incident CHD among Black Americans, including 4-hydroxyphenylacetate, phenylacetyl-L-glutamine, imidazole-propionate, 3-hydroxy-2-ethylpropionate, trans-4-hydroxyproline, and TMAO (all OR>1.25 in quantitative stage). The associations between circulating microbial metabolites and health outcomes may vary among individuals with different ethnicities and dietary/lifestyle habits, or from different regions (thus, different environmental exposures). For example, TMAO was significant among Americans but not among Chinese, possibly due to varied dietary sources of TMAO precursors (e.g., red meat vs. fish).^42,44^ We also found stronger associations of 3-hydroxy-2-ethylpropionate and 3-hydroxybutyrate among individuals without obesity, suggesting impaired BCAA and ketone body catabolism may underlie “metabolically unhealthy normal weight” phenotype and CVD risk. Further research on these pathways may be fruitful. Yet, since most metabolites showed no significant p-interaction by race, future larger studies with diverse populations are needed to evaluate if certain metabolites are population-specific or general CVD risk factors.

Several limitations of our current study merit discussion. First, we were not able to validate all significant metabolites in the discovery or in-silico validation. Due to differences in metabolomics assays, 12 significant metabolites from discovery were unavailable for in-silico validation, including butyrate, l-urobilinogen, and an unnamed metabolite (X-12411, associated with incident CHD in EPIC-Norfolk). Also, given the limited coverage of our quantitative assay, 16 metabolites that passed in-silico validation could not be measured simultaneously, such as dihomo-linolenate (20:3n3 or n6) and sphingomyelin (d18:1/21:0, d17:1/22:0, d16:1/23:0). Specific assays for these metabolites are needed to verify the findings. Second, although we broadly adjusted for potential confounders, including sociodemographics, lifestyles, and history of metabolic diseases, residual confounding may still exist due to inadequate adjustment or unmeasured confounders. Third, despite validation in independent samples, the causal role of highlighted microbial metabolites in CHD etiology and therapeutics requires investigation. Emerging studies manipulating the gut microbiome or metabolites show promise for CVD prevention and treatment.^4,45^ Finally, none of the cohorts collected stool samples at blood draw, so we could not link microbiome or fecal metabolites with blood metabolites. However, circulating metabolites reflect functional outputs of the microbiome and microbiota-host interactions most relevant to the host cardiovascular system, which also represent the most feasible way to study gut microbial metabolism with incident CHD in existing large cohorts. Future studies collecting multiple types of biospecimens, following participants for incident CVD, and applying multi-omics will further shed light on the role of microbiota-host interactions in CVD development and prevention.

In conclusion, we systematically examined the associations between circulating gut microbiota-related metabolites and incident CHD among demographically and geographically diverse participants. We identified and validated microbial metabolites of amino acids, lipids, and xenobiotics linked to incident CHD. Our findings support the important role of gut microbiota and microbial metabolism in CHD etiology and highlight promising microbial metabolites and pathways that may serve as novel biomarkers or therapeutic targets for future mechanistic and interventional studies.

## Supporting information

Supplemental Methods, Fig, and Ref

## Data Availability

Data underlying the article are available upon reasonable request and approval by the Data Use Committees of the participating cohorts with a signed Data Use Agreement.

## Acknowledgment

The authors thank the staff and participants of the SCCS, SWHS, SMHS, MESA, and ARIC study for their important contributions.

## Author contributions

D.Y. designed the study and drafted the manuscript. Y.Z., J.J.Y., G.S., N.Q.N., and R.P. analyzed the data. D.Y., D.K.G., L.L., D.H., B.Y., I.T., H.C., Q.C., X.O.S, and W.Z. contributed to obtaining the data and funding. All authors contributed to the interpretation of the results and critically reviewed and edited the manuscript.

## Data Availability Statement

The data underlying this article can be obtained through the Southern Community Cohort Study (https://www.southerncommunitystudy.org) and Shanghai Women’s Health Study & Shanghai Men’s Health Study (https://swhs-smhs.app.vumc.org/index.php) upon reasonable request with approval by the cohort committees and a signed data use agreement.

## Supplementary Materials

Supplementary Methods, Supplementary Figure 1, Supplementary Tables 1-4

## References

1. Martin SS, Aday AW, Allen NB, et al. 2025 Heart Disease and Stroke Statistics: A Report of US and Global Data From the American Heart Association. Circulation. Jan 27 2025;doi:10.1161/CIR.0000000000001303

2. Lindstrom M, DeCleene N, Dorsey H, et al. Global Burden of Cardiovascular Diseases and Risks Collaboration, 1990-2021. J Am Coll Cardiol. Dec 20 2022;80(25):2372–2425. doi:10.1016/j.jacc.2022.11.001

3. Aron-Wisnewsky J, Clement K. The gut microbiome, diet, and links to cardiometabolic and chronic disorders. Nat Rev Nephrol. Mar 2016;12(3):169–81. doi:10.1038/nrneph.2015.191

4. Tang WHW, Hazen SL. Unraveling the Complex Relationship Between Gut Microbiome and Cardiovascular Diseases. Circulation. May 14 2024;149(20):1543–1545. doi:10.1161/CIRCULATIONAHA.123.067547

5. Valles-Colomer M, Menni C, Berry SE, Valdes AM, Spector TD, Segata N. Cardiometabolic health, diet and the gut microbiome: a meta-omics perspective. Nat Med. Mar 2023;29(3):551–561. doi:10.1038/s41591-023-02260-4

6. Witkowski M, Weeks TL, Hazen SL. Gut Microbiota and Cardiovascular Disease. Circ Res. Jul 31 2020;127(4):553-570. doi:10.1161/CIRCRESAHA.120.316242

7. Takeuchi T, Nakanishi Y, Ohno H. Microbial Metabolites and Gut Immunology. Annu Rev Immunol. Jun 2024;42(1):153-178. doi:10.1146/annurev-immunol-090222-102035

8. Roessler J, Leistner DM, Landmesser U, Haghikia A. Modulatory role of gut microbiota in cholesterol and glucose metabolism: Potential implications for atherosclerotic cardiovascular disease. Atherosclerosis. Oct 2022;359:1–12. doi:10.1016/j.atherosclerosis.2022.08.018

9. Krautkramer KA, Fan J, Backhed F. Gut microbial metabolites as multi-kingdom intermediates. Nat Rev Microbiol. Feb 2021;19(2):77–94. doi:10.1038/s41579-020-0438-4

10. Agus A, Clement K, Sokol H. Gut microbiota-derived metabolites as central regulators in metabolic disorders. Gut. Jun 2021;70(6):1174–1182. doi:10.1136/gutjnl-2020-323071

11. Nightingale Health Biobank Collaborative G. Metabolomic and genomic prediction of common diseases in 700,217 participants in three national biobanks. Nat Commun. Nov 21 2024;15(1):10092. doi:10.1038/s41467-024-54357-0

12. Pietzner M, Stewart ID, Raffler J, et al. Plasma metabolites to profile pathways in noncommunicable disease multimorbidity. Nat Med. Mar 2021;27(3):471–479. doi:10.1038/s41591-021-01266-0

13. Nogal A, Alkis T, Lee Y, et al. Predictive metabolites for incident myocardial infarction: a two-step meta-analysis of individual patient data from six cohorts comprising 7897 individuals from the COnsortium of METabolomics Studies. Cardiovasc Res. Dec 30 2023;119(17):2743–2754. doi:10.1093/cvr/cvad147

14. Fromentin S, Forslund SK, Chechi K, et al. Microbiome and metabolome features of the cardiometabolic disease spectrum. Nat Med. Feb 2022;28(2):303–314. doi:10.1038/s41591-022-01688-4

15. Wang Z, Zhu C, Nambi V, et al. Metabolomic Pattern Predicts Incident Coronary Heart Disease. Arterioscler Thromb Vasc Biol. Jul 2019;39(7):1475–1482. doi:10.1161/ATVBAHA.118.312236

16. Cruz DE, Tahir UA, Hu J, et al. Metabolomic Analysis of Coronary Heart Disease in an African American Cohort From the Jackson Heart Study. JAMA Cardiol. Feb 1 2022;7(2):184–194. doi:10.1001/jamacardio.2021.4925

17. Wu GD, Compher C, Chen EZ, et al. Comparative metabolomics in vegans and omnivores reveal constraints on diet-dependent gut microbiota metabolite production. Gut. Jan 2016;65(1):63–72. doi:10.1136/gutjnl-2014-308209

18. Walejko JM, Kim S, Goel R, et al. Gut microbiota and serum metabolite differences in African Americans and White Americans with high blood pressure. Int J Cardiol. Nov 15 2018;271:336–339. doi:10.1016/j.ijcard.2018.04.074

19. De Filippis F, Pellegrini N, Vannini L, et al. High-level adherence to a Mediterranean diet beneficially impacts the gut microbiota and associated metabolome. Gut. Nov 2016;65(11):1812–1821. doi:10.1136/gutjnl-2015-309957

20. Signorello LB, Hargreaves MK, Blot WJ. The Southern Community Cohort Study: investigating health disparities. J Health Care Poor Underserved. Feb 2010;21(1 Suppl):26-37. doi:10.1353/hpu.0.0245

21. Zheng W, Chow WH, Yang G, et al. The Shanghai Women’s Health Study: rationale, study design, and baseline characteristics. Am J Epidemiol. Dec 1 2005;162(11):1123–31. doi:10.1093/aje/kwi322

22. Shu XO, Li H, Yang G, et al. Cohort Profile: The Shanghai Men’s Health Study. Int J Epidemiol. Jun 2015;44(3):810–8. doi:10.1093/ije/dyv013

23. Wright JD, Folsom AR, Coresh J, et al. The ARIC (Atherosclerosis Risk In Communities) Study: JACC Focus Seminar 3/8. J Am Coll Cardiol. Jun 15 2021;77(23):2939–2959. doi:10.1016/j.jacc.2021.04.035

24. The Atherosclerosis Risk in Communities (ARIC) Study: design and objectives. The ARIC investigators. Am J Epidemiol. Apr 1989;129(4):687–702.

25. Bild DE, Bluemke DA, Burke GL, et al. Multi-Ethnic Study of Atherosclerosis: objectives and design. Am J Epidemiol. Nov 1 2002;156(9):871–81.

26. Evans AM, DeHaven CD, Barrett T, Mitchell M, Milgram E. Integrated, nontargeted ultrahigh performance liquid chromatography/electrospray ionization tandem mass spectrometry platform for the identification and relative quantification of the small-molecule complement of biological systems. Anal Chem. Aug 15 2009;81(16):6656–67. doi:10.1021/ac901536h

27. Neveu V, Nicolas G, Amara A, Salek RM, Scalbert A. The human microbial exposome: expanding the Exposome-Explorer database with gut microbial metabolites. Sci Rep. Feb 2 2023;13(1):1946. doi:10.1038/s41598-022-26366-w

28. Dona AC, Jimenez B, Schafer H, et al. Precision high-throughput proton NMR spectroscopy of human urine, serum, and plasma for large-scale metabolic phenotyping. Anal Chem. Oct 7 2014;86(19):9887–94. doi:10.1021/ac5025039

29. Tzoulaki I, Castagne R, Boulange CL, et al. Serum metabolic signatures of coronary and carotid atherosclerosis and subsequent cardiovascular disease. Eur Heart J. Sep 7 2019;40(34):2883–2896. doi:10.1093/eurheartj/ehz235

30. Lewis MR, Pearce JT, Spagou K, et al. Development and Application of Ultra-Performance Liquid Chromatography-TOF MS for Precision Large Scale Urinary Metabolic Phenotyping. Anal Chem. Sep 20 2016;88(18):9004–13. doi:10.1021/acs.analchem.6b01481

31. Nemet I, Li XS, Haghikia A, et al. Atlas of gut microbe-derived products from aromatic amino acids and risk of cardiovascular morbidity and mortality. Eur Heart J. Aug 22 2023;44(32):3085–3096. doi:10.1093/eurheartj/ehad333

32. Ottosson F, Brunkwall L, Smith E, et al. The gut microbiota-related metabolite phenylacetylglutamine associates with increased risk of incident coronary artery disease. J Hypertens. Dec 2020;38(12):2427–2434. doi:10.1097/HJH.0000000000002569

33. Qi Q, Li J, Yu B, et al. Host and gut microbial tryptophan metabolism and type 2 diabetes: an integrative analysis of host genetics, diet, gut microbiome and circulating metabolites in cohort studies. Gut. Jun 2022;71(6):1095–1105. doi:10.1136/gutjnl-2021-324053

34. de Mello VD, Paananen J, Lindstrom J, et al. Indolepropionic acid and novel lipid metabolites are associated with a lower risk of type 2 diabetes in the Finnish Diabetes Prevention Study. Sci Rep. Apr 11 2017;7:46337. doi:10.1038/srep46337

35. Wang Z, Peters BA, Yu B, et al. Gut Microbiota and Blood Metabolites Related to Fiber Intake and Type 2 Diabetes. Circ Res. Mar 29 2024;134(7):842–854. doi:10.1161/CIRCRESAHA.123.323634

36. Ryan RO. Metabolic annotation of 2-ethylhydracrylic acid. Clin Chim Acta. Aug 25 2015;448:91–7. doi:10.1016/j.cca.2015.06.012

37. Tobias DK, Lawler PR, Harada PH, et al. Circulating Branched-Chain Amino Acids and Incident Cardiovascular Disease in a Prospective Cohort of US Women. Circ Genom Precis Med. Apr 2018;11(4):e002157. doi:10.1161/CIRCGEN.118.002157

38. Ruiz-Canela M, Toledo E, Clish CB, et al. Plasma Branched-Chain Amino Acids and Incident Cardiovascular Disease in the PREDIMED Trial. Clin Chem. Apr 2016;62(4):582–92. doi:10.1373/clinchem.2015.251710

39. Fine KS, Wilkins JT, Sawicki KT. Circulating Branched Chain Amino Acids and Cardiometabolic Disease. J Am Heart Assoc. Apr 2 2024;13(7):e031617. doi:10.1161/JAHA.123.031617

40. Tang WH, Wang Z, Levison BS, et al. Intestinal microbial metabolism of phosphatidylcholine and cardiovascular risk. N Engl J Med. Apr 25 2013;368(17):1575–84. doi:10.1056/NEJMoa1109400

41. Koeth RA, Wang Z, Levison BS, et al. Intestinal microbiota metabolism of L-carnitine, a nutrient in red meat, promotes atherosclerosis. Nat Med. May 2013;19(5):576–85. doi:10.1038/nm.3145

42. Yang JJ, Shu XO, Herrington DM, et al. Circulating trimethylamine N-oxide in association with diet and cardiometabolic biomarkers: an international pooled analysis. Am J Clin Nutr. May 8 2021;113(5):1145–1156. doi:10.1093/ajcn/nqaa430

43. Lopaschuk GD, Dyck JRB. Ketones and the cardiovascular system. Nat Cardiovasc Res. May 2023;2(5):425-437. doi:10.1038/s44161-023-00259-1

44. Landfald B, Valeur J, Berstad A, Raa J. Microbial trimethylamine-N-oxide as a disease marker: something fishy? Microb Ecol Health Dis. 2017;28(1):1327309. doi:10.1080/16512235.2017.1327309

45. Li F, Armet AM, Korpela K, et al. Cardiometabolic benefits of a non-industrialized-type diet are linked to gut microbiome modulation. Cell. Jan 20 2025;doi:10.1016/j.cell.2024.12.034

